# The importance of saturating density dependence for predicting SARS-CoV-2 resurgence

**DOI:** 10.1101/2020.08.28.20183921

**Authors:** E. S. Nightingale, O. J. Brady, CMMID Covid-19 working group, L. Yakob

## Abstract

Severe acute respiratory syndrome coronavirus 2 (SARS-CoV-2) associated mortality data from England show evidence for an increasing trend with population density until a saturating level, after adjusting for local age distribution, deprivation, proportion of ethnic minority population and proportion of key workers among the working population. Projections from a mathematical model that accounts for this observation deviate markedly from the current status quo for SARS-CoV-2 models which either assume linearity between density and transmission (30% of models) or no relationship at all (70%). Respectively, these standard model structures over- and under-estimate the delay in infection resurgence following the release of lockdown. Identifying saturation points for given populations and including transmission terms that account for this feature will improve model accuracy and utility for the current and future pandemics.

## Introduction

Like many pathogens that cause respiratory diseases (*1-3*), severe acute respiratory syndrome coronavirus 2 (SARS-CoV-2) appears to be transmitted more effectively in densely populated areas (*4-6*). The increased disease rates reported among high-density populations (*4, 5, 7, 8*) may, however, be an artefact of confounders, such as the higher proportion of individuals of lower socioeconomic status or from minority ethnic groups in urban areas (*9*). Using COVID-19 associated mortality data from the Office for National Statistics, we aimed to assess the evidence for density dependence.

Standard transmission models that either do or do not account for this density dependence have been used interchangeably because their projections are generally equivalent when population density remains unperturbed or is homogeneous, e.g. at a national level. While the ~1% infection fatality rate for COVID-19 (*10*) is insufficient to destabilize populations, the reaction of most countries’ governments to curtail disease spread through mass quarantine (‘lockdown’) and social distancing has had unprecedented impacts on the density of mobile human populations. For example, the UK’s lockdown, which came into effect on March 23^rd^ 2020, effectively reduced the freely moving population from 66.5 million to 10.6 million (key workers) (*11*). This same intervention was employed by numerous countries, similarly impacting their mobile populations (*12*). We evaluate the extent to which models built to inform the epidemiology of COVID-19 use an underlying structure that can accommodate the drastic changes and variation in densities experienced by most global populations.

As lockdowns are gradually released over the latter part of 2020, global populations will re-equilibrate to a ‘new normal’ whereby densities of mobile people are increased but in which contact patterns are expected to remain reduced through social distancing interventions (*13*). Using a suite of mathematical models we illustrate the impact that the different, routinely ignored, assumptions underlying transmission and density may have in projecting infection dynamics and measuring intervention effectiveness.

## Results

### Evidence for saturating density dependence in COVID-19 associated deaths

COVID-19 associated deaths appear to be strongly correlated with population density, aligning with the rural/urban disparities demonstrated in Office for National Statistics bulletins (Supplementary Fig 1, (*14*)). Adjusting for potential confounders (age distribution, deprivation, ethnic distribution, proportion of key workers within the local population) via a negative binomial generalized linear model a saturating dependence on population density provided the best fit to total local authority mortality rates over the period March 3rd to July 31st with respect to the leave-one-out information criterion (LOOIC) (Fig 1a). Models independent of, or linearly dependent on, density performed similarly since the fitted linear trend was negligibly small, and both performed worse than log-linear and saturating forms. The chosen model suggests a 3.6- fold (90% CrI [2.44, 5.28]) increase in mortality rate for a standard deviation difference in density, on the saturated scale. Owing to the heightened risk of earlier outbreak seeding for higher density areas, we repeated the analysis additionally adjusting for the lag of the local epidemic behind the national. The saturating model was retained as the best fit (Table 1) and suggested a similar increase in mortality rate of 4-fold (90% CrI [2.15, 7.08]).

**Fig 1.**
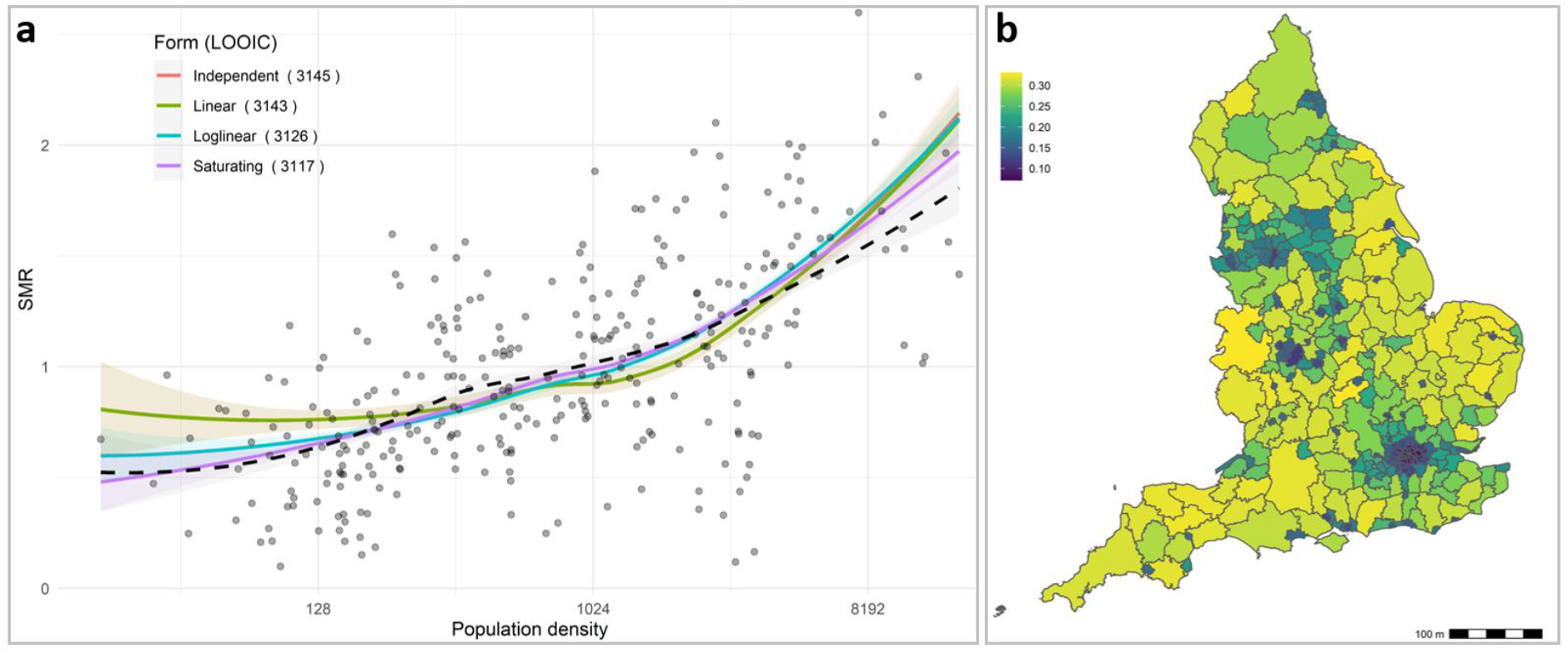
Dependence of observed versus age-specific expected mortality rates (standardized mortality ratio, SMR) on population density. **a)** Four forms of density dependence (and loess curve, dashed black line) are illustrated in the left panel, with LOOIC values for each fit demonstrating superiority of the saturating density-dependent function. **b)** The heterogeneous impact of 84% effective density reduction on the proportional reduction in predicted mortality among the freely moving population according to the saturating model is mapped in the right panel.

**Table 1.** Model comparison for explaining variation in mortality rates. Models additionally account for the local epidemic lags behind the national and are compared on LOOIC, with all compared to the optimal model (saturating form) in the first row. Saturating and log-linear forms are not clearly distinguishable from each other, but both appear preferable over the independent and linear forms.

|  | LOOIC | LOOIC SE | Difference elpd | Diff. elpd SE |
| --- | --- | --- | --- | --- |
| Saturating | 1904 | 22.7 | 0.0 | 0.0 |
| Log-linear | 1908 | 22.4 | -2.4 | 1.0 |
| Independent | 1923 | 21.6 | -9.6 | 4.5 |
| Linear | 1924.1 | 20.6 | -10.3 | 4.4 |

Under the saturating density-dependent model, the impact of lockdown on reducing transmission among mobile individuals, and consequently deaths, is heterogeneous - having greatest benefit to regions with low population density (>30% reduction in projected deaths for example in Devon, Herefordshire and the Derbyshire Dales) but reduced benefit to high-density regions (~5-7% reduction for the London boroughs of Tower Hamlets, Hackney, Islington and Camden) (Fig 1b). These results were retained when accounting for the lag of the local epidemic behind the national (Supplementary Fig 2).

### Projecting SARS-CoV-2 resurgence after lockdown is released

A full text review of 100 epidemiological models of SARS-CoV-2 showed that 70% explicitly assume that contact rate between people (and, hence transmission) is unaffected by population density (Supporting Information). Of the remaining 30% of models, all assumed a linear relationship between population density and transmission.

We use a metapopulation model to simulate the infection dynamics among freely moving as well as locked-down individuals, incorporating transmission terms that can accommodate density-independent (referred to as ‘frequency-dependent’) as well as linearly and saturating density-dependent assumptions. We show that while all functional forms perform equivalently in fitting mortality data leading up to lockdown, dynamics under alternate assumptions may diverge markedly during and following the phase when lockdown is released (Fig. 2). We note that any adaptive public health responses (i.e. additional interventions) curbing the second wave are ignored - this comparison is intended to illustrate the consequences to projected dynamics of alternative assumptions underlying density and transmission.

**Fig. 2.**
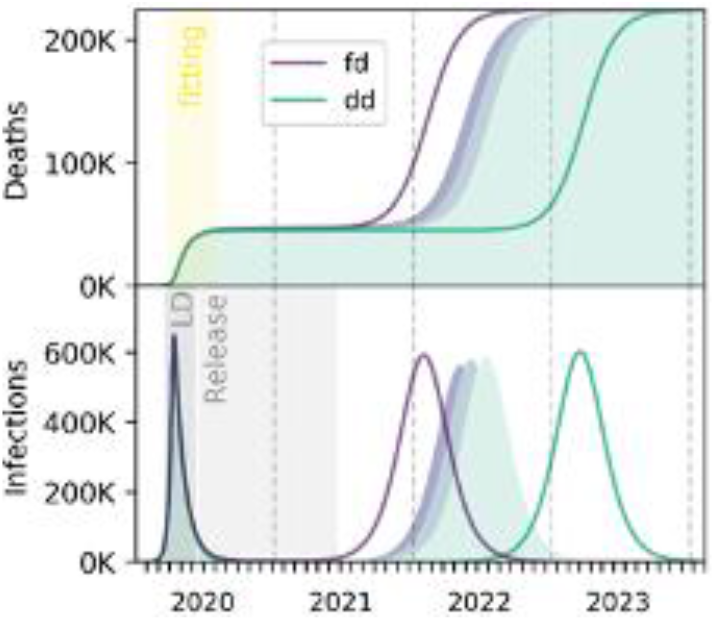
Population density and SARS-CoV-2 dynamics. COVID-19 associated mortality (top) and infection dynamics (bottom) following the release of lockdown ‘LD’ under the three different transmission terms (frequency-dependent ‘fd’, linearly density-dependent ‘dd’ and saturating density-dependent). Lighter filled areas illustrate saturating density-dependent dynamics for lower population density (where England’s density is set to equal that of London at 5700 people per Km^2^, the average English population density at 430 people per Km^2^ or Cornwall at 160 people per Km^2^). These simulations show a 1-year release of locked down individuals and infection preventative behaviors (e.g. face masks) that halve the per contact transmission rate. Details of model-fitting are in the supplementary materials.

Although final epidemic size and total deaths were equivalent for the alternative classic assumptions, transmission was delayed by over a year under a density-versus frequency-dependent model (Fig 2). This delay occurs because only under the density-dependent assumption the force of infection is reduced while any part of the population remains locked down. At the very high densities of London populations, locking down 84% of people under our saturating density-dependent model had an impact most similar to a frequency-dependent assumption. Meaning, if the density of England’s entire population was equivalent to the density found in London, infection dynamics and deaths resulting from a saturating density-dependent model most closely match the frequency-dependent projections (although, with a 3-month lag). However, London has a population density that is an order of magnitude higher than the next most populated region in England; and projected infection dynamics diverged more considerably under scenarios reflecting densities experienced outside of the capital. The force of infection and the timing of peak prevalence for the saturating density-dependent model is constrained between the frequency-and linearly density-dependent versions (*15*) with lower densities tending towards the latter.

Assuming a maximum national capacity of 5000 intensive care unit (ICU) beds, we then assess the difference between these temporal limits in the projected duration between the release of lockdown and a second wave of infection exceeding ICU capacity (Fig. 3). Threshold levels of social distancing to interrupt transmission (i.e., maintain effective reproduction number below one) are similar for both classic models. However, where interventions fail to achieve this threshold, density-dependent transmission resulted in a delay of a year before ICU inundation. This is contingent on the timeframe across which lockdown is released, whereby more gradual releases extend delays.

**Fig. 3.**
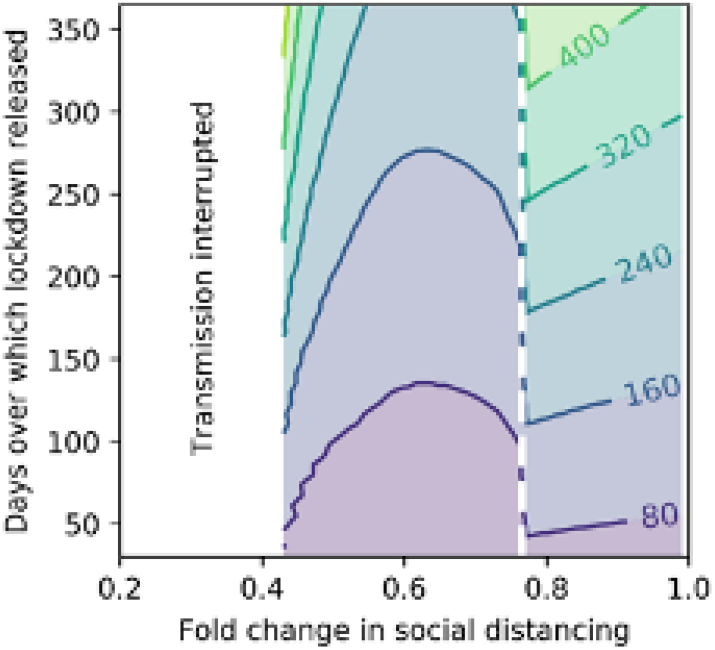
Consequences of density dependence on intensive care unit inundation. Difference (in days, contours) between models in their projected time before ICU capacity is exceeded, as a function of lockdown release schedule and effectiveness of social distancing. The dashed white line marks minimum social distancing required to prevent immediate ICU inundation under the frequency-dependent model.

## Discussion

Projections of COVID-19 infection dynamics following the release of a huge proportion of the population from lockdown comprise an urgent and critical component of public health decision making (*16*). The classical forms of modelling infectious diseases among populations have been used interchangeably by different research groups because, under most plausible circumstances, they exhibit equivalent dynamics. In March 2020, England locked down over four-fifths of its population. For most, this fundamentally altered the rate at which people made contact. Under the current circumstance of millions of people easing out of lockdown, substantial differences between projections from a frequency- and density-dependent transmission assumption emerge. Most notably, density dependence results in delayed infection resurgence; and, contingent on the timeframe across which lockdown is released and the effectiveness of social distancing, this delay can extend to over a year.

The delay is a function of a fundamental aspect of density-dependent transmission: lower host densities reduce the force of infection, and there is a threshold host density below which an infection cannot spread. Despite its origins in human infectious disease modelling (*17*), the existence of this threshold has historically had limited epidemiological application. The phenomenon is discussed more widely in wildlife disease ecology (*18*) where it underlies key disease-control decisions such as culling (*19*). Current expectation is that lockdowns, either full or of a more moderate or localized form, will be reimplemented when cases restart increasing. Density effects and thresholds are particularly pertinent in the current pandemic during which extreme fluctuations in mobile human density are likely to continue.

Analyzing COVID-19 associated deaths across different regions in England, and accounting for known major confounders (*9*), the non-linear increase in deaths with population density was adequately captured by neither classical form of modelling transmission. Using a function that captures the saturating increase in deaths with population density resulted in an expedited resurgence compared with a linearly density-dependent model and a delayed resurgence compared with the popularly used frequency-dependent model.

Less populated areas were shown to have fewer per capita deaths (as per England’s mortality data) and slower resurgences following the release of lockdown. This provides more achievable targets and considerably more lead time for health services to prepare than would otherwise be anticipated. It also highlights a hazard. During and after releases from lockdown, in order to fit a prolonged lag in cases, transmission rates derived from most current (frequency-dependent) models will underestimate the effective reproduction number. This could exaggerate the perceived effectiveness of ongoing interventions, such as social distancing or face masks, with potentially serious consequences.

Our study is limited by the fact that we do not have comprehensive data on how contact rates were affected prior to and over the lockdown period for individuals inhabiting regions of differing population density. We also do not know where people were infected, only where they were when they died. Instead we have had to resort to mortality rates and locations as a proxy. It is possible, for example, that contact rates are not affected by dramatic shifts in population density regardless of baseline levels (i.e., the average England resident came into contact with as many individuals during lockdown as prior to lockdown, satisfying a frequency-dependent assumption), and that the increased per capita fatality seen in more densely populated regions has an alternative, thus far unidentified explanation. Mobile phone applications developed to inform participants of urgent health information have already gained millions of users in the current coronavirus context (*20*). Piggy-backing on these efforts could help substantiate the evidence for the contact-density relationships we have identified.

Due to the highly complex interactions between population characteristics, behaviors and mortality risk, the association discovered between saturating density and mortality rates may remain confounded by factors not considered here. Moreover, the criterion used for model comparison depends on an independence assumption which may not hold between neighboring LTLAs. Work is ongoing to characterize the patterns of spatial correlation in mortality at the LTLA level.

Infectious diseases are emerging at an unprecedented rate (*21*) and the upwards trend in global travel and urbanization increases the likelihood of pandemics (*22*). Their success in controlling SARS-CoV-2 means that widescale lockdowns will not only continue to be enforced as this pandemic progresses, but they will likely be more readily applied in future emergencies. It is crucial that we use the current opportunity to collect data to inform more precise forms of how contact rates are altered at varying stages of lockdown. Future work should also address whether the feature we have identified from England’s data is generalizable to other countries. Incorporating realistic contact-density relationships into the transmission term of population-level mathematical models will improve precision of their projections and their utility in public health decision making.

## Materials and Methods

### Experimental Design

- A statistical analysis of COVID-19 associated mortality in England at the lower-tier local authority level.
- A review of SARS-CoV-2 mathematical models (see Supplementary materials).
- A mathematical modelling analysis of the consequence of density-dependent SARS-CoV-2 transmission to post-lockdown resurgence.

### Data

Reported COVID-19-related deaths between the 01-03-2020 and 31-07-2020 were obtained in anonymized linelist form from Public Health England, and were filtered to include all deaths which occurred within 28 days of positive COVID-19 test (N = 36,311). Individual records were aggregated to lower-tier local authority (LTLA), and nationally by 10-year age bands in order to calculate age-standardized expected counts.

Local authority shapefiles and single-age population estimates were obtained from ONS (*23*). Four sub-regions of Buckinghamshire (Aylesbury Vale, Chiltern, South Bucks, Wycombe) were aggregated in order to match most recent population estimates. The City of London was aggregated with Westminster due to its very small resident population, and the Isles of Scilly excluded since no COVID-19-related deaths had been reported there during this period. Index of multiple deprivation (IMD) (*24*), percentage of minority ethnic population (*25*) and percentage of key workers among the working population (*26*) are characteristics of the LTLA population potentially associated with both COVID-19 mortality and population density, therefore were included as covariates in all models. Percentage of key workers was missing for Westminster and Cornwall; these were imputed by the median value across all neighboring LTLAs.

### Statistical analysis

Negative binomial regression models were fit to the number of deaths (n) per LTLA, adjusting for the four covariates (IMD, % minority population, % key workers and lag in weeks behind the first death nationally). Age distribution within the LTLA was adjusted for via inclusion of age-adjusted expected deaths (*E*) as an offset; these were calculated according to national age-specific rates (deaths per 100,000 per age band) applied to local population estimates in ten-year age bands. Population density was accounted for in one of four functional forms:

A. Constant/independent of population density
B. Linear
C. Log-linear
D. Saturating

This yields the following model specification:

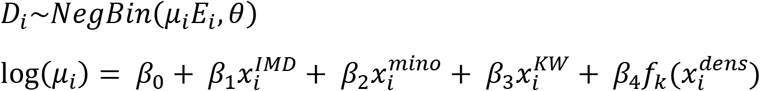

where

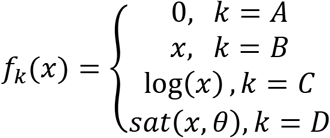

and

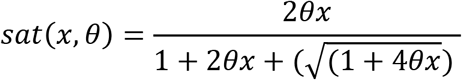

Models were fitted using the *rstanarm* package (*27*) with default weakly-informative priors. The four model variants were compared on LOOIC, calculated via approximate leave-one-out cross validation as implemented in the *loo* package (*28*). Interpretation is the same as that of the AIC in that smaller values reflect better fit. The value of *θ* for the saturating function was determined by manual optimization of the glm with respect to LOOIC on a hold-out set of 40% of LTLAs, over a range from 0.001 to 1.

For the saturating model, the impact of an 84% reduction in effective population density as a result of lockdown on predicted mortality rates among the freely moving population was calculated as a percentage change between mean model-predicted deaths under the original and reduced densities.

### Mathematical model

We use a discrete-time, deterministic compartmental model (Fig. 4) with daily timesteps to simulate SARS-CoV-2 transmission. From the first day of lockdown (March 23^rd^ 2020), 84% of the population enter quarantine in which frequency-dependent transmission occurs. This assumption is made for the lockdown sub-population because an individual’s likelihood of contracting infection while in their home is limited by their household size (i.e., not impacted by the density of individuals under quarantine in different households). Each model is fitted independently to England’s COVID-19 associated mortality data (up until August 1^st^ 2020). We compare frequency-dependent and both linearly and saturating density-dependent transmission among the remaining free-movers for when lockdown is released. We also explore the impact of varying rates of migration between locked down and free-moving individuals (Supplementary Fig 3). We compare frequency-dependent and linearly density-dependent transmission (the limiting cases for the saturating density-dependent model (*15*)) among the remaining free-movers for a range of lockdown release schedules (over a period of between 1 and 12 months). Contact rates are reduced through two distinct mechanisms under the density-dependent models: whereas reduced contact through social distancing behavioral changes among the freely moving population (e.g. the 2-metre rule) is included in all models, only the density-dependent versions assume reduced opportunities for mobile people coming into contact with others because of their substantially depleted numbers. Full model specification and sources for its parameterization can be found in the Supplementary Materials, and the Python (v3.8) code is freely available from https://github.com/lwyakob/COVIDsaturates.

**Fig. 4.**
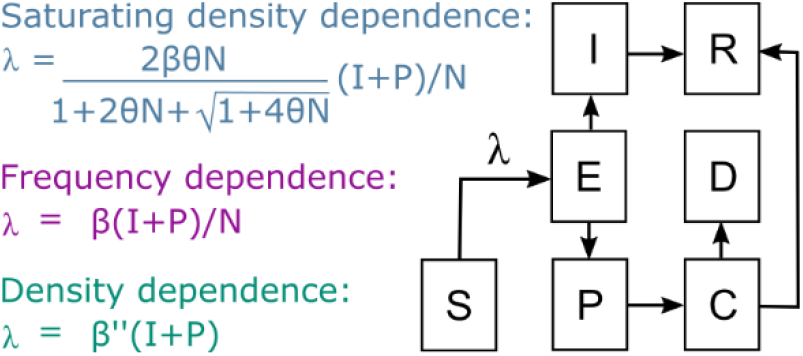
Model compartments and alternative transmission assumptions. Model compartments are: ‘S’usceptible, ‘E’xposed, ‘I’nfectious, ‘P’re-critical infectious, ‘C’ritically ill, ‘D’ead and ‘R’ecovered. Under a frequency-dependent assumption, the force of infection, λ, is the product of the transmission coefficient, β, and the proportion of the total population, N, that are infectious. Under a density-dependent assumption, the force of infection is the product of the transmission coefficient and the density of infectious individuals. The saturating density-dependent formulation assumes the force of infection is a product of the transmission coefficient and a function of the density of infectious individuals and parameter *θ* derived from analyzing England’s regional mortality data.

## Data Availability

All data can be retrieved from our github

https://github.com/lwyakob/COVIDsaturates

## Funding

OJB was funded by a Sir Henry Wellcome Fellowship funded by the Wellcome Trust (206471/Z/17/Z). ESN is funded by the Bill and Melinda Gates Foundation (OPP1183986).

## Author contributions

LY conceived of the study, reviewed COVID-19 models and performed the mathematical modelling analysis. EN and OJB performed the statistical analysis. All authors contributed towards results interpretations and writing.

## Competing interests

There are no competing interests.

## Supplementary Materials

### Supplementary Materials and Methods

Google Scholar search by year ‘2020’ on June 19th 2020:

**coronavirus, OR covid19, OR covid-19 “‘(mathematical OR simulation OR transmission) model’“**

613 results were returned and ordered by relevance. These were consecutively reviewed to ascertain if they used SIR-related models and whether they declared (either explicitly stating or in presented equations) which functional form of transmission was used in their analysis, until 100 relevant articles were found for inclusion in this review. The following are the list of papers sorted by whether they use linearly density- or frequency-dependent transmission.

Linearly density-dependent transmission
1. Aleta, A., et al., Modeling the impact of social distancing, testing, contact tracing and household quarantine on second-wave scenarios of the COVID-19 epidemic. medRxiv, 2020.
2. Arenas, A., et al., A mathematical model for the spatiotemporal epidemic spreading of COVID19. MedRxiv, 2020.
3. Bentout, S., A. Chekroun, and T. Kuniya, Parameter estimation and prediction for coronavirus disease outbreak 2019 (COVID-19) in Algeria. AIMS Public Health, 2020. 7(2): p. 306.
4. Boudrioua, M.S. and A. Boudrioua, Predicting the COVID-19 epidemic in Algeria using the SIR model. medRxiv, 2020.
5. Chatterjee, K., et al., Healthcare impact of COVID-19 epidemic in India: A stochastic mathematical model. Medical Journal Armed Forces India, 2020. 76(2): p. 147-55.
6. Diop, B.Z., et al., The relatively young and rural population may limit the spread and severity of Covid-19 in Africa: a modelling study. BMJ global health, 2020. 5(5): p. e002699.
7. Eguiluz, V.M., et al., Risk of secondary infection waves of COVID-19 in an insular region: the case of the Balearic Islands, Spain. medRxiv, 2020.
8. Engbert, R., et al., Sequential data assimilation of the stochastic SEIR epidemic model for regional COVID-19 dynamics. medRxiv, 2020.
9. Giordano, G., et al., A SIDARTHE model of COVID-19 epidemic in Italy. arXiv, 2020.
10. González, R.E., C. Estupiñán-López, and C.A.C. Morales, An adapted ODE model to study the Dynamics of SARS-Cov-2 Infection (COVID-19): different scenarios for Brazil and other countries. Research Square, 2020.
11. Guinet, A., A modelling of COVID-19 outbreak with a linear compartmental model. HAL, 2020.
12. Kissler, S.M., et al., Projecting the transmission dynamics of SARS-CoV-2 through the postpandemic period. Science, 2020. 368(6493): p. 860-868.
13. Mbabazi, F.K., et al., A Mathematical Model Approach for Prevention and Intervention Measures of the COVID-19 Pandemic in Uganda. MedRxiv, 2020.
14. Ng, K.Y. and M.M. Gui, COVID-19: Development of a robust mathematical model and simulation package with consideration for ageing population and time delay for control action and resusceptibility. Physica D: Nonlinear Phenomena, 2020: p. 132599.
15. Pai, C., A. Bhaskar, and V. Rawoot, Investigating the dynamics of COVID-19 pandemic in India under lockdown. Chaos, Solitons & Fractals, 2020. 138: p. 109988.
16. Pais, R.J. and N. Taveira, Predicting the evolution and control of the COVID-19 pandemic in Portugal. F1000Research, 2020. 9(283): p. 283.
17. Pedro, S.A., et al., Conditions for a second wave of COVID-19 due to interactions between disease dynamics and social processes. medRxiv, 2020.
18. Peirlinck, M., et al., Outbreak dynamics of COVID-19 in China and the United States. Biomechanics and modeling in mechanobiology, 2020: p. 1-15.
19. Pellis, L., et al., Challenges in control of Covid-19: short doubling time and long delay to effect of interventions. arXiv, 2020.
20. Prem, K., et al., The effect of control strategies to reduce social mixing on outcomes of the COVID-19 epidemic in Wuhan, China: a modelling study. The Lancet Public Health, 2020. 5(5): p. E261-E270.
21. Prieto, F., et al., COVID-19 Impact Estimation on ICU Capacity at Andalusia, Spain, Using Artificial Intelligence. Research Square, 2020.
22. Rasha, M. and S. Balamuralitharan, A study on COVID-19 transmission dynamics: Stability analysis of SEIR model with Hopf bifurcation for effect of time delay. Research Square, 2020.
23. Saha, S. and S. Saha, The impact of the undetected COVID-19 cases on its transmission dynamics. medRxiv, 2020.
24. Sarma, U. and B. Ghosh, Quantitative modeling and analysis show country-specific optimization of quarantine measures can potentially circumvent COVID19 infection spread post lockdown. medRxiv, 2020.
25. Schwartz, I.B., et al., Predicting the impact of asymptomatic transmission, non-pharmaceutical intervention and testing on the spread of COVID19 COVID19. medRxiv, 2020.
26. Wahid, A., et al., The Epidemiology of COVID-19 and applying Non Pharmaceutical interventions by using the Susceptible, Infectious Recovered epidemiological Model in Pakistan. medRxiv, 2020.
27. Wickramaarachchi, T. and S. Perera, Optimal Control Measures to Combat COVID 19 Spread in Sri Lanka: A Mathematical Model Considering the Heterogeneity of Cases. medRxiv, 2020.
28. Wickramaarachchi, T., S. Perera, and S. Jayasinghe, COVID-19 epidemic in Sri Lanka: A mathematical and computational modelling approach to control. medRxiv, 2020.
29. Yafia, R., Modeling and Dynamics in Epidemiology, COVID19 with Lockdown and Isolation Effect: Application to Moroccan Case. medRxiv, 2020.
30. Zongo, P., et al., A model of covid-19 transmission to understand the effectiveness of the containment measures: application to French data. HAL, 2020.

Frequency-dependent transmission
1. Abrigo, M.R., et al., Projected Disease Transmission, Health System Requirements, and Macroeconomic Impacts of the Coronavirus Disease 2019 (COVID-19) in the Philippines. 2020, Discussion Paper Series.
2. Al-Shammari, A.A., et al., Real-time tracking and forecasting of the COVID-19 outbreak in Kuwait: a mathematical modeling study. medRxiv, 2020.
3. Ali, M., M. Imran, and A. Khan, Can medication mitigate the need for a strict lock down?: A mathematical study of control strategies for COVID-19 infection. medRxiv, 2020.
4. Ali, M., et al., The role of asymptomatic class, quarantine and isolation in the transmission of COVID-19. Journal of Biological Dynamics, 2020. 14(1): p. 389-408.
5. Alshammari, F.S., A mathematical model to investigate the transmission of COVID-19 in the Kingdom of Saudi Arabia. medRxiv, 2020.
6. Armstrong, E., M. Runge, and J. Gerardin, Identifying the measurements required to estimate rates of COVID-19 transmission, infection, and detection, using variational data assimilation. MedRXiv, 2020.
7. Assob, J.-C., D. Dongo, and P.E. Nguimkeu, Early Dynamics of Transmission and Projections of COVID-19 in Some West African Countries. 2020.
8. Atkeson, A., What will be the economic impact of COVID-19 in the US? Rough estimates of disease scenarios. 2020, National Bureau of Economic Research.
9. Ayoub, H.H., et al., Characterizing key attributes of the epidemiology of COVID-19 in China: Model-based estimations. medRxiv, 2020.
10. Bherwani, H., et al., Exploring Dependence of COVID-19 on Environmental Factors and Spread Prediction in India. 2020.
11. Blasius, B., Power-law distribution in the number of confirmed COVID-19 cases. arXiv, 2020.
12. Brugnago, E.L., et al., How relevant is the decision of containment measures against COVID-19 applied ahead of time? arXiv, 2020.
13. Casella, F., Can the COVID-19 epidemic be managed on the basis of daily data? arXiv, 2020.
14. Chen, X. and Z. Qiu, Scenario analysis of non-pharmaceutical interventions on global COVID-19 transmissions. arXiv, 2020.
15. Chudik, A., M.H. Pesaran, and A. Rebucci, Voluntary and mandatory social distancing: Evidence on covid-19 exposure rates from chinese provinces and selected countries. 2020, National Bureau of Economic Research.
16. Cuevas, J.A.G., SEI1I2HRSVM model applied to the coronavirus pandemic (COVID-19) in Paraguay. arXiv, 2020.
17. Davies, N.G., et al., Age-dependent effects in the transmission and control of COVID-19 epidemics. Nature Medicine, 2020.
18. Davies, N.G., et al., Effects of non-pharmaceutical interventions on COVID-19 cases, deaths, and demand for hospital services in the UK: a modelling study. The Lancet Public Health, 2020.
19. Davies, N.G., et al., The impact of Coronavirus disease 2019 (COVID-19) on health systems and household resources in Africa and South Asia. medRxiv, 2020.
20. Dehning, J., et al., Inferring change points in the spread of COVID-19 reveals the effectiveness of interventions. Science, 2020.
21. Duczmal, L.H., et al., Vertical social distancing policy (‘Isolamento Vertical’) is ineffective to contain the coronavirus COVID-19 pandemic. Cadernos de Saúde Pública, 2020.
22. Dziugys, A., et al., Simplified model of Covid-19 epidemic prognosis under quarantine and estimation of quarantine effectiveness. medRxiv, 2020.
23. Eikenberry, S.E., et al., To mask or not to mask: Modeling the potential for face mask use by the general public to curtail the COVID-19 pandemic. Infectious Disease Modelling, 2020. 5: p. 293-308.
24. Francis, A., et al., Projected ICU and Mortuary load due to COVID-19 in Sydney. medRxiv, 2020.
25. Gatto, M., et al., Spread and dynamics of the COVID-19 epidemic in Italy: Effects of emergency containment measures. Proceedings of the National Academy of Sciences, 2020. 117(19): p. 10484-10491.
26. Gupta, M., et al., Transmission dynamics of the COVID-19 epidemic in India and modelling optimal lockdown exit strategies. medRxiv, 2020.
27. Harris, J.E., The Coronavirus Epidemic Curve is Already Flattening in New York City. 2020, National Bureau of Economic Research.
28. Hasan, A., et al., A new estimation method for COVID-19 time-varying reproduction number using active cases. arXiv, 2020.
29. Hochberg, M.E., Importance of suppression and mitigation measures in managing COVID-19 outbreaks. arXiv, 2020.
30. Iboi, E.A., et al., Mathematical Modeling and Analysis of COVID-19 pandemic in Nigeria. medRxiv, 2020.
31. Ivorra, B., et al., Mathematical modeling of the spread of the coronavirus disease 2019 (COVID-19) taking into account the undetected infections. The case of China. Communications in nonlinear science and numerical simulation, 2020. 88: p. 105303.
32. Jitsuk, N.C., et al., Effect of the Songkran festival on COVID-19 transmission in Thailand. Asian Pacific Journal of Tropical Medicine, 2020. 13(7): p. 331-332.
33. Kassa, S.M., J.B. Njagarah, and Y.A. Terefe, Analysis of the mitigation strategies for COVID-19: from mathematical modelling perspective. Chaos, Solitons & Fractals, 2020. 138: p. 109968.
34. Ke, R., et al., Fast spread of COVID-19 in Europe and the US suggests the necessity of early, strong and comprehensive interventions. medRxiv, 2020.
35. Kennedy, D.M., et al., Modeling the effects of intervention strategies on COVID-19 transmission dynamics. Journal of Clinical Virology, 2020. 128: p. 104440.
36. Khailaie, S., et al., Estimate of the development of the epidemic reproduction number Rt from Coronavirus SARS-CoV-2 case data and implications for political measures based on prognostics. medRxiv, 2020.
37. Khajanchi, S., et al., Dynamics of the COVID-19 pandemic in India. arXiv, 2020.
38. Kouakep, Y., et al., Modelling the anti-COVID19 individual or collective containment strategies in Cameroon, in Epi-Ndere. 2020: At University of Ngaoundere (Cameroon).
39. Kumar, S., S. Sharma, and N. Kumari, Future of COVID-19 in Italy: A mathematical perspective. arXiv, 2020.
40. Kuzdeuov, A., et al., A Network-Based Stochastic Epidemic Simulator: Controlling COVID-19 with Region-Specific Policies. medRxiv, 2020.
41. Kyagulanyi, A., et al., Risk analysis and prediction for COVID19 demographics in low resource settings using a python desktop app and excel models. medRxiv, 2020.
42. Leng, T., et al., The effectiveness of social bubbles as part of a Covid-19 lockdown exit strategy, a modelling study. medRxiv, 2020.
43. López, L. and X. Rodo, A modified SEIR model to predict the COVID-19 outbreak in Spain and Italy: simulating control scenarios and multi-scale epidemics. Available at SSRN 3576802, 2020.
44. Luo, J., Predictive Monitoring of COVID-19. SUTD Data-Driven Innovation Lab, 2020.
45. Lyra, W., et al., COVID-19 pandemics modeling with SEIR (+ CAQH), social distancing, and age stratification. The effect of vertical confinement and release in Brazil. medRxiv, 2020.
46. MANOU-ABI, S. and J. BALICCHI, Analysis of the COVID-19 epidemic in french overseas department Mayotte based on a modified deterministic and stochastic SEIR model. medRxiv, 2020.
47. Mukaddes, A.M.M. and M. Sannyal, Transmission Dynamics of COVID-19 in Bangladesh-A Compartmental Modeling Approach. Research Square, 2020.
48. Neto, O.P., et al., COVID-19 mathematical model reopening scenarios for Sao Paulo-Brazil. medRxiv, 2020.
49. Nyabadza, F., et al., Modelling the potential impact of social distancing on the COVID-19 epidemic in South Africa. medRxiv, 2020.
50. Ogbunugafor, C.B., et al., Variation in SARS-CoV-2 free-living survival and environmental transmission can modulate the intensity of COVID-19 outbreaks. medRxiv, 2020.
51. Pant, R., et al., COVID-19 Epidemic Dynamics and Population Projections from Early Days of Case Reporting in a 40 million population from Southern India. medRxiv, 2020.
52. Picchiotti, N., et al., COVID-19 Italian and Europe epidemic evolution: A SEIR model with lockdown-dependent transmission rate based on Chinese data. Available at SSRN 3562452, 2020.
53. Picchiotti, N., et al., COVID-19 pandemic: a mobility-dependent SEIR model with undetected cases in Italy, Europe and US. arXiv, 2020.
54. Prabhakaran, H., Spread of the Novel Coronavirus (SARS-CoV-2): Modeling and Simulation of Control Strategies. medRxiv, 2020.
55. Rahman, M.M., et al., Impact of control strategies on COVID-19 pandemic and the SIR model based forecasting in Bangladesh. medRxiv, 2020.
56. Rajesh, A., et al., CoVID-19 prediction for India from the existing data and SIR (D) model study. medRxiv, 2020.
57. Rapolu, T., et al., A Time-Dependent SEIRD Model for Forecasting the COVID-19 Transmission Dynamics. medRxiv, 2020.
58. Rawson, T., et al., How and When to End the COVID-19 Lockdown: An Optimization Approach. Frontiers in Public Health, 2020. 8: p. 262.
59. Reis, R.F., et al., Characterization of the COVID-19 pandemic and the impact of uncertainties, mitigation strategies, and underreporting of cases in South Korea, Italy, and Brazil. Chaos, Solitons & Fractals, 2020: p. 109888.
60. Rouabah, M.T., A. Tounsi, and N.-E. Belaloui, Early dynamics of COVID-19 in Algeria: a model-based study. arXiv, 2020.
61. Salomon, J.A., Defining high-value information for COVID-19 decision-making. medRxiv, 2020.
62. Sardar, T., S.S. Nadim, and J. Chattopadhyay, Assessment of 21 days lockdown effect in some states and overall India: a predictive mathematical study on COVID-19 outbreak. arXiv, 2020.
63. Tuite, A.R., D.N. Fisman, and A.L. Greer, Mathematical modelling of COVID-19 transmission and mitigation strategies in the population of Ontario, Canada. CMAJ, 2020. 192(19): p. E497-E505.
64. Tuite, A.R., et al., Risk for COVID-19 Resurgence Related to Duration and Effectiveness of Physical Distancing in Ontario, Canada. Annals of Internal Medicine, 2020(M20-2945).
65. Verachi, F., L.G. Trussoni, and L. Lanzi, CoViD-19 in Italy: a mathematical model to analyze the epidemic containment strategy and the economic impacts. medRxiv, 2020.
66. Xu, R., et al., Weather Conditions and COVID-19 Transmission: Estimates and Projections. Available at SSRN 3593879, 2020.
67. Yang, P., et al., The effect of multiple interventions to balance healthcare demand for controlling COVID-19 outbreaks: a modelling study. medRxiv, 2020.
68. Zhang, J., et al., Changes in contact patterns shape the dynamics of the COVID-19 outbreak in China. Science, 2020.
69. Zhang, Y., et al., Applicability of time fractional derivative models for simulating the dynamics and mitigation scenarios of COVID-19. Chaos, Solitons & Fractals, 2020: p. 109959.
70. Matrajt, L. and T. Leung, Early Release-Evaluating the Effectiveness of Social Distancing Interventions to Delay or Flatten the Epidemic Curve of Coronavirus Disease. Emerging Infectious Diseases, 2020. 26(8).

### Mathematical model equations

Free-moving subpopulation:

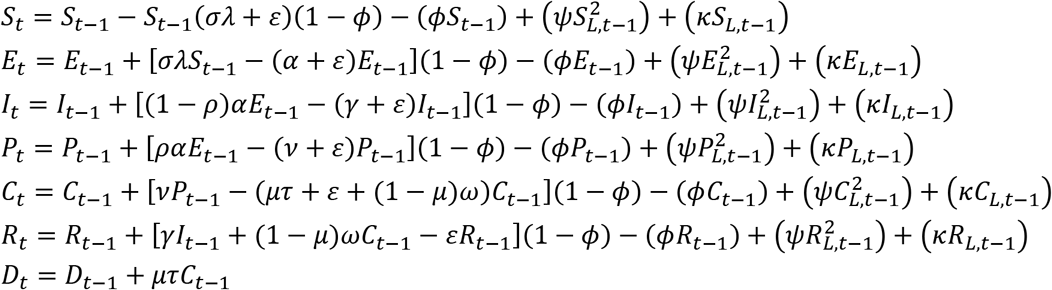

Locked down subpopulation:

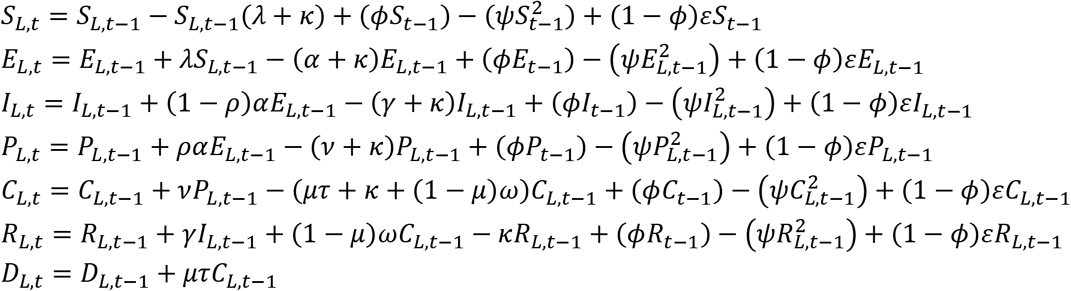

The transmission rate in the absence of any intervention was set assuming an R_0_ of 3 (*28*). Data captured deaths pre-lockdown, during lockdown and just following the initial release of individuals after lockdown. During lockdown, the population split into two sub-populations. The transmission rates were generated by fitting deaths in the model to England’s mortality data retrieved from (*29*). At the same time as lockdown, social distancing also reduced per capita contact rates among the free-moving population:

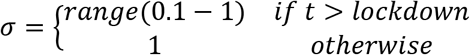

For the full range of social distancing, the model was refitted to the mortality data (more stringent social distancing among free-movers requiring less of a reduction in the lockdown transmission rate, β_L_). Least squares fitting using the Levenberg-Marquardt minimization algorithm was conducted using ‘lmfit’ in Python v3.8.

## Supplementary figures

**Fig S1.**
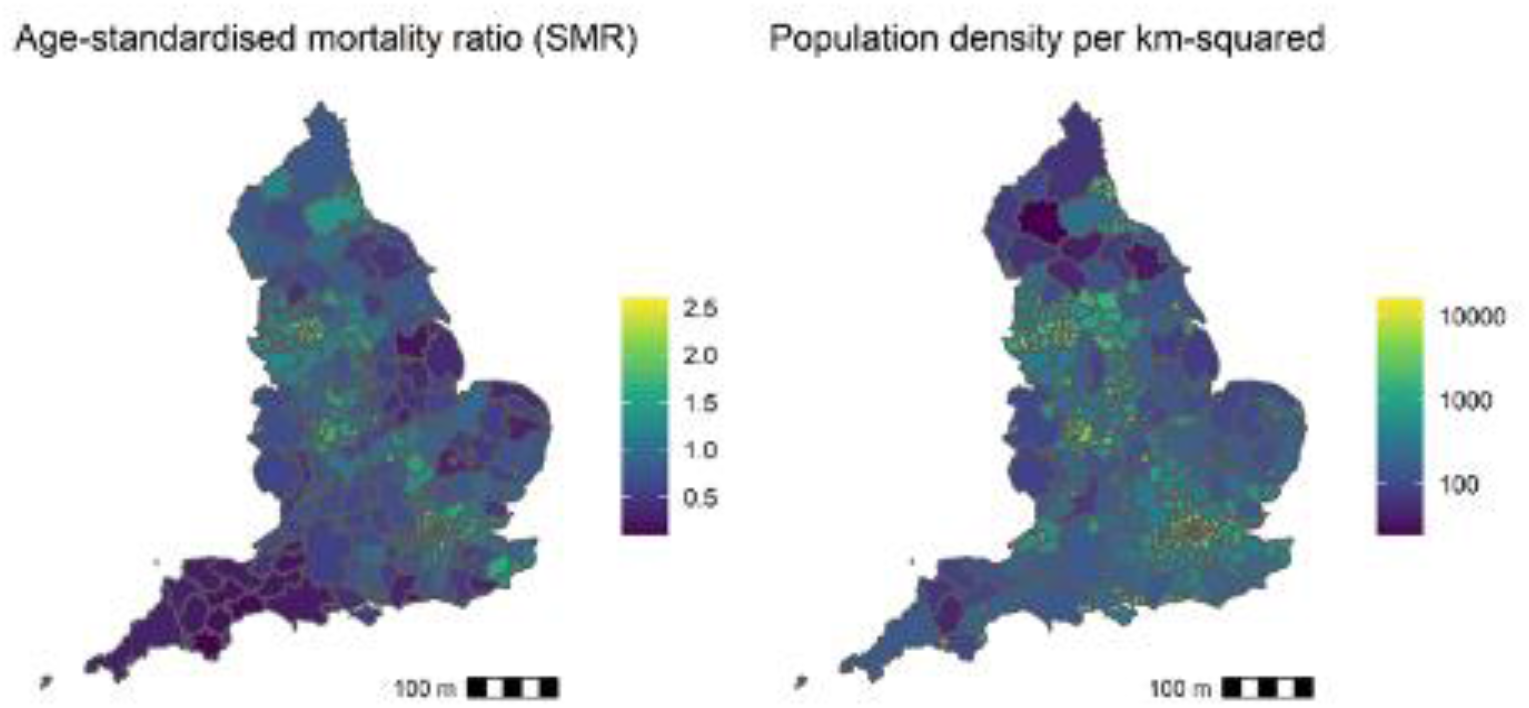
COVID-19 associated mortality and population density in England. Age-standardized mortality ratios are mapped in the left panel and population density per km^2^ in the right, by lower-tier local authority.

**Fig S2.**
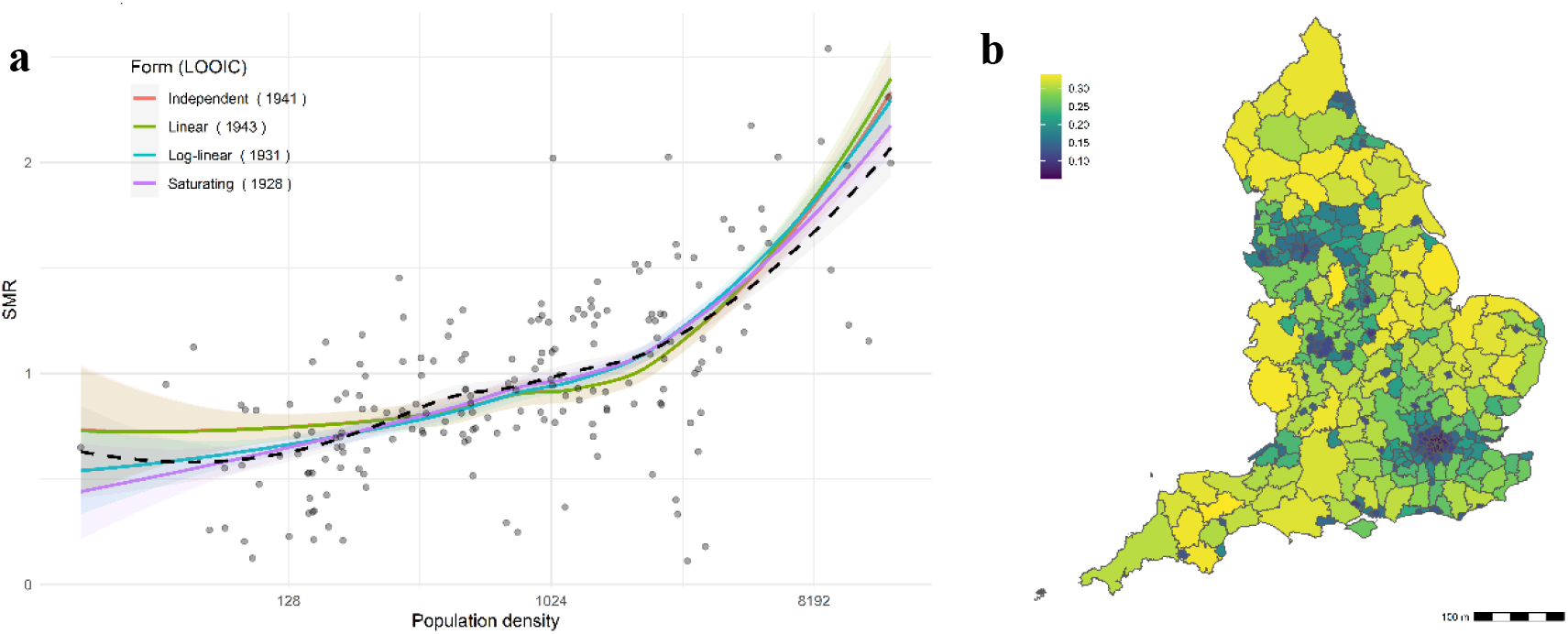
Dependence of observed versus age-specific expected mortality rates (standardized mortality ratio, SMR) on population density. The effects of different outbreak seed timings for different LTLAs are incorporated. **a)** Four forms of density dependence (and loess curve, dashed black line) are illustrated in the left panel, with LOOIC values for each fit demonstrating superiority of the saturating density-dependent function. **b)** The heterogeneous impact of 84% effective density reduction on the proportional reduction in predicted mortality among the freely moving population according to the saturating model is mapped in the right panel.

**Fig. S3.**
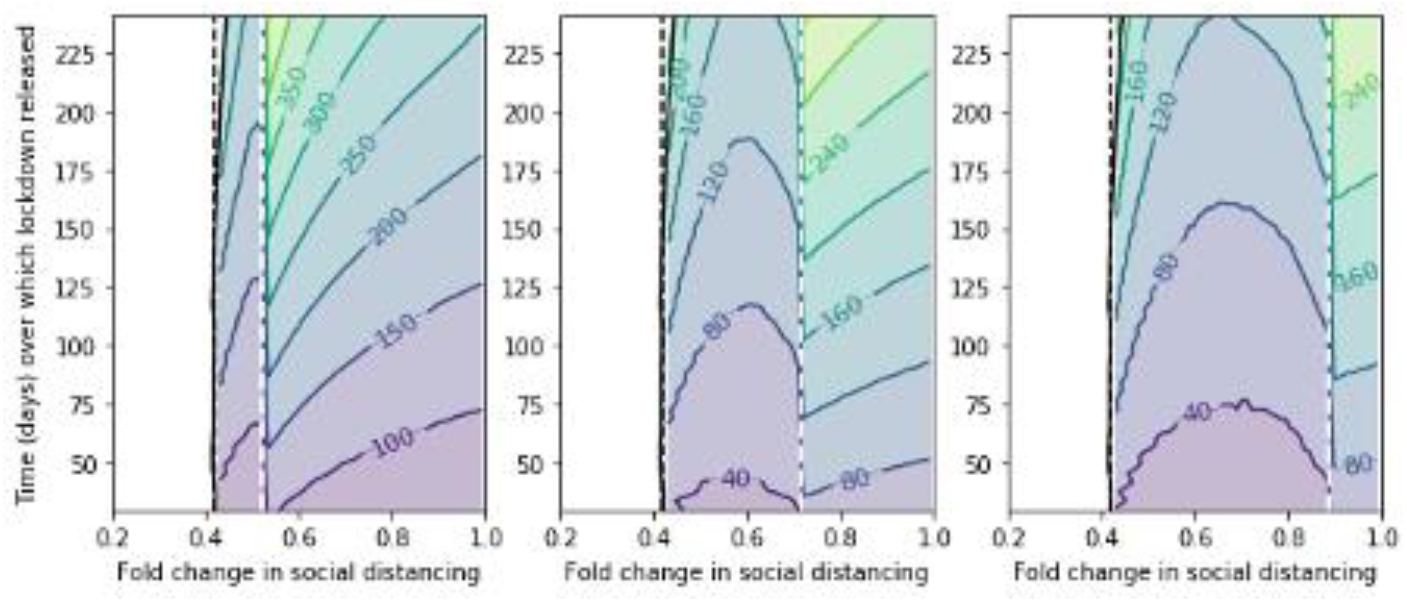
Sensitivity analysis of movements between sub-populations. The difference between functional forms in projected time (contours=days) until the ICU capacity is exceeded by critically ill patients. Left: 50% movement (‘*ε*’), Middle: 100% *ε*, Right: 150% *ε*. For each value of *ε* the model was refitted to the data (the density-dependent model was insensitive to movement; for the frequency-dependent model, higher rates of movement required slightly less of a reduction in transmission among the lockdown sub-population). Black lines mark thresholds for interrupting transmission for density-independent (dashed) and -dependent (solid) models. Dashed white lines mark minimum social distancing required to prevent immediate ICU inundation under the frequency-dependent model.

## Supplementary tables

**Table S1.** Mathematical model variables

| Variable | Definition |
| --- | --- |
| $S_t, S_{L,t}$ | Susceptible population at time 't'; subscript 'L' denotes lockdown sub-population |
| $E_t, E_{L,t}$ | Exposed population at time 't' |
| $I_t, I_{L,t}$ | Infected population at time 't' |
| $P_t, P_{L,t}$ | Pre-critically infected population at time 't' |
| $C_t, C_{L,t}$ | Critically infected population at time 't' |
| $R_t, R_{L,t}$ | Recovered population at time 't' |
| $D_t, D_{L,t}$ | Dead population at time 't' |

**Table S2.** Mathematical model parameters

| Parameter | Definition | Value | Source |
| --- | --- | --- | --- |
| $\lambda$ | Force of infection (composite of other parameters and variables) | n/a | n/a |
| $\beta''$ | Transmission rate (linearly density-dependent assumption) | | derived |
| $\beta$ | Transmission rate | | derived |
| $\beta_L$ | Transmission rate while under lockdown (frequency dependent) | | derived |
| $\varepsilon$ | Daily rate of movement from free-moving to lockdown sub-pop | 0.1 but see sensitivity analysis section | |
| $\kappa$ | Daily rate of movement from lockdown to free-moving sub-pop | | |
| $\phi$ | Pulsed, mass movement of 84% free-movers into lockdown | 0.84 | (11) |
| $\psi$ | Daily rate of release of lockdown sub-pop | Wide range tested | |
| $\sigma$ | Fold change in social contacts among free-movers | Wide range tested | |
| $\alpha$ | Inverse of infection latent period | 1/5.8 | (31) |
| $\rho$ | Proportion of infected individuals becoming critically infected | 0.02 | (32) |
| $\gamma$ | Inverse of recovery period | 1/5 | (33) |
| $\nu$ | Inverse of additional delay before symptoms become critical | 1/7 | (34) |
| $\mu$ | Proportion of critically infected that die | 0.427 | (35) |
| $\tau$ | Inverse of time for critically ill to die | 1/7 | (36) |
| $\omega$ | Inverse of time for critically ill to recover | 1/7.2 | (35) |

## Notes

### Competing Interest Statement

The authors have declared no competing interest.

### Funding Statement

OJB was funded by a Sir Henry Wellcome Fellowship funded by the Wellcome Trust (206471/Z/17/Z). Neither the authors nor their institutions at any time received payment or services from a third party for any aspect of the submitted work.

### Summary of Updates

We added to the statistical analysis to include LTLA-level lags in seeded outbreaks relative to the national. The main results and conclusions remain unchanged following this inclusion.

